# Teledentistry for improving access to, and quality of oral health care: A protocol for an overview of systematic reviews and meta-analyses

**DOI:** 10.1101/2023.07.04.23292218

**Authors:** Pascaline Kengne Talla, Paul Allison, André Bussières, Nicolas Giraudeau, Svetlana Komarova, Quentin Basiren, Frédéric Bergeron, Elham Emami

## Abstract

Digital technologies are becoming essential to address and optimize the suboptimal performance of healthcare systems. Teledentistry involves the use of information and communication technology to improve access to oral health care and the quality of oral health care delivery. Several systematic reviews (SRs) have been conducted to synthesize evidence on the effectiveness of teledentistry but with conflicting results. The aim of this review is to comprehensively summarize available SRs and provide evidence on the impact of teledentistry on access to oral care, patients’ and oral healthcare providers’ outcomes, quality of oral health care and costs. This protocol has been registered with the International Prospective Register of Systematic Reviews (PROSPERO CRD42022373964). Six electronic databases including MEDLINE (Ovid), Embase (Embase.com), CINAHL (EBSCO), Web of Science, Cochrane Library and Epistemonikos will be searched for SRs of quantitative, qualitative, and mixed reviews evaluating teledentistry modalities involving both patients and/or oral health care providers (OHCPs). We will include only studies published in English or French. The primary outcomes will be considered from the patients’ perspective (e.g., access to oral health care, patient-reported outcomes, and experiences). The secondary outcomes will include outcomes from patients and OHCPs (e.g., clinical outcomes, safety, behaviors, and costs). Two independent reviewers will perform data screening, data extraction and will assess the quality of included studies using the AMSTAR 2 and ROBIS tools. Data will be synthesized narratively and presented by tables and graphs. We will report any overlap of primary studies in the SRs. A statement on the strength of evidence for each outcome will be provided if possible. This review will inform decision-makers, patients, OHCPs, and researchers on the potential effectiveness, benefits, and challenges of teledentistry and support them in making recommendations for its use. Results will be disseminated through peer-reviewed publications, presentations at conferences, and on social media.

## Introduction

Oral diseases globally affect more than 3.5 billion people, highlighting the need for interventions that could improve accessibility to and affordability of oral health care [1]. Information and communication technologies (ICTs) are promising approaches to address some of the inadequacies of healthcare systems, to improve patients’ access to and experiences of care, to reduce the costs of care delivery and to promote high value care [2–8]. In addition, digital technologies can improve the quality of health care (e.g equity, safety, effectiveness, patient-centered and timely care) [9, 10]. The COVID-19 pandemic has accelerated the use of these technologies in all health-related disciplines including dentistry [11], paving the way for the development of virtual dental care. They have reshaped the delivery of oral health care including patient-clinician interaction, screening, diagnosis of oral diseases, monitoring of patients, treatment planning and management of care [12].

Emerging evidence suggests that teledentistry, a branch of telehealth, is cost-effective at the micro, meso and macro levels involving patients, oral health care providers (OHCPs) and allied health care workers, and decision makers [13–16]. Teledentistry includes synchronous and asynchronous modes, and uses different vehicles such as telephones, smartphones, tablets and computers, as well as various approaches such as calls, text and voice messaging, videos, and applications [17, 18]. Teledentistry has been used in the context of dental education, training, and transfer of information or for the delivery of dental care/services. It involves the interaction between OHCPs and their peers, or with other health care providers or with their patients and/or caregivers to improve patients’ outcomes and the quality of care.

Multiple reviews have synthesized the evidence on the benefits and implementation challenges of teledentistry [19–22], the process of teledentistry, and the outcomes and experiences [13, 14, 23–25] from the perspective of patients, healthcare organizations and OHCPs [26]. However, the results are often conflicting [22, 27, 28], inconsistent or inconclusive [21, 29, 30]. Although systematic reviews are a compelling means of synthesizing research, a systematic review of existing systematic reviews (SRs) can provide a broader assessment of the quality and credibility of available evidence [31, 32], and offer valuable information for patients, families, health professionals, researchers, and policy-makers. The information generated by such overview can be used to enhance both clinical practice and population health. In this study, we will use the term ‘overview’ due to the lack of consistency in the literature on the terminology of the compilation data from multiple SRs to provide a single summary of relevant evidence [33, 34]. A published overview on teledentistry evaluated its accuracy and effectiveness for the delivery of oral health care [35]. A major limitation of this overview is the lack of risk of bias assessments of the included SRs. Moreover, it has only focused on accuracy of screening, diagnosis, and therapeutic management of dental care outcomes. Other health outcomes related to access to oral care (e.g., utilization of services) and patients’ and OHCPs’ behaviors to improve the quality of oral healthcare would be important to consider to inform practice, policy decision-making, and future research [36]. Another available overview is limited to tele-orthodontics to improve compliance in orthodontic patients [37]. Therefore, there is a need to address the gaps in literature by conducting a comprehensive overview of existing SRs using a rigorous methodology with valid quality assessment of the evidence [38]. This proposed overview of existing SRs aims to compile and contrast the existing evidence from systematic reviews published on teledentistry [31]. Accurate information resulting from this overview will assist decision-makers on the effectiveness of teledentistry and inform the development of guidelines to support OHCPs in its implementation.

### Research question

We will answer to the following research question: “From the perspective of a range of stakeholders, to what extent is teledentistry effective in improving access to, and quality of oral healthcare, while reducing related costs?”

## Methods

The review is guided by the Joanna Briggs Institute (JBI) Manual for Evidence synthesis for umbrella reviews [39], and the Cochrane handbook for overview of reviews [40], and is written in accordance with Preferred Reporting Items for Systematic Reviews and Meta-Analyses Protocols (PRISMA-P) guidance [41]. The PRISMA checklist is available as S1 checklist.

### Study registration

This overview has been registered in the International Prospective Register of Systematic Reviews (PROSPERO CRD42022373964).

### Eligibility criteria

We will use the “PICOSS” format: Participants, Intervention, Comparator, Outcome, Study design and Setting.

#### Participants/Population

We will include SRs involving patients receiving oral healthcare services performed by any licensed OHCPs (general dental practitioners, dental specialists, dental hygienists, dental nurses/assistants, and dental therapists) and the SRs involving OHCPs with or without the patients.

#### Intervention

Teledentistry refers to use of information and communication technologies including the transmission of clinical information and images between an oral health professional and patient or between two health professionals, including at least one oral health professional, who are separated by distance for dental consultations, diagnosis and treatment planning [18]. Teledentistry is a modality used to provide remote access to healthcare services to patients. It includes the use of a group of technologies and modalities, which can be categorized as follows [17]: i) Store and forward, which is used to keep patients’ oral health records such as radiographs and photographs; ii) Remote patient monitoring, used for patient data collection from a remote site and then transferring them to a dental practitioner in another location; iii) Live video, involving the use of a real-time interaction between a patient and an OHCP, using audio-visual communication for screening, diagnosis, treatment planning or follow-up; and iv) Mobile health, making use of mobile communication devices such as phones and tablets to provide virtual oral healthcare services. Other terms referring to teledentistry encompass e-health, virtual care, telemedicine oral health and mobile oral health. Irrespective of the term used, we will include all interventions that involve oral health care delivery through telecommunication systems in the presence of at least one member of dental staff.

#### Comparator

We will include any SRs where the comparator could be one of the following types of interventions or cases: in-person or face-to face interventions or usual care; no intervention; synchronous versus asynchronous; synchronous versus mobile health; synchronous versus remote monitoring; and other digital technologies (e.g. electronic dental records, virtual reality).

#### Outcomes

The primary outcomes are reported from the patients’ perspective. They will include access to oral health care (e.g. use of oral healthcare services, number of consultations, use in emergency cases, delay of treatment, waiting time); patient-reported outcomes (e.g. oral health related- and overall quality of life; self-reported clinical outcomes; pain management, oral functions; psychosocial impact) and experiences with oral health care (e.g. satisfaction with care; communication with OHCPs, patient-centered care and empowerment; acceptance and understanding of information and confidence in the treatment, and experience with the technology such as ability to use the application).

Secondary outcomes will be reported as they relate to:

- Patient indicators: Clinical outcomes reported by OHCPs (e.g. plaque index, gingival index and white spot lesions); Adherence/compliance to treatment (e.g. medication, oral health prevention and promotion practices); Knowledge, attitude, and behavior; Barriers and enablers towards the use of teledentistry; Safety; Adverse outcomes; and Costs (e.g. travel time, transportation, missing work/school, loss of the productivity and consultation time).
- OHCP indicators: Accuracy of diagnosis; Awareness, knowledge, attitude, and behavior; Barriers and enablers towards the adoption of teledentistry; Prescribing (e.g. testing, medication); Monitoring of patients; Coordination and management of oral health care; Communication with patients, dental specialists, dental staff and other health professionals; Costs (e.g. equipment, number of patients per day, number of consultations per day, waiting time, training in the use of the equipment for teledentistry); and Equity.

#### Study designs

We will include systematic reviews with or without meta-analysis of quantitative (randomized control trials, quasi-experimental studies such as non-randomized controlled trials, before and after controlled studies and interrupted series studies, or observational studies), qualitative and mixed methods studies using any teledentistry modalities (asynchronous, remote monitoring, real-time, and mhealth). The term SR refers to a reproducible, standardized and transparent approach aiming to identify, evaluate and summarize the evidence from primary studies on a particular topic, thereby making it more accessible to decision-makers [42]. We will include only SRs that have conducted searches in at least two databases, have clear inclusion/exclusion criteria, performed quality assessment, and synthesized included studies [32, 42, 43].

#### Setting

We will capture the evidence from SRs on teledentistry conducted in any dental setting (e.g., dental offices, school, community, hospital, home care), geographical region (e.g., rural, urban), and country (low-, middle- or high income).

### Exclusion criteria

We will exclude any types of knowledge synthesis systematic reviews lacking a formal methodological quality or risk of bias assessment or where a search was conducted in a single database (26). We will exclude studies on teledentistry focusing only on education and training in dentistry and research without a care delivery component.

### Search strategy

We will search electronic databases in MEDLINE (Ovid), Embase, CINAHL (EBSCO), Web of Science, The Cochrane Library and Epistemonikos (https://www.epistemonikos.org/) using a search strategy that has been developed using an interactive process by members of the research team with the support of an expert librarian. The search strategy will be conducted from database inception using the following keywords: (“teledentistry” OR “remote care*” OR “mobile health”) AND (“reviews” OR “meta-analysis” OR “systematic review). We will also perform a search in Sociological Abstract (Proquest), Academic Search Premier (EBSCO), and Proquest Dissertations & Theses. We will contact experts in the field by email if additional data are required. In addition, we will check the reference lists of included SRs and any identified overviews for any eligible articles. See S1 file for the search strategy conducted in Medline. We will update the search prior to the publication of the review to identify any new relevant systematic reviews. There will be no restrictions for countries, age of participants, publication date and settings. However, we will consider only studies published in English or French because of the limited resources for translation.

### Study selection

SRs identified by the search will be imported into Covidence software [44]. The research team will discuss the selection criteria to ensure a shared understanding before pairs of assessors will independently pilot the initial screening phase (i.e., titles and abstracts) on 10% of total number of citations retrieved. Disagreements at each stage will be resolved through discussion or consultation with a third reviewer whenever necessary. We will adjust the grid following this calibration exercise. Reviewers will consider “include”, “exclude”, or “Maybe” as modalities of the studies selection. All studies rated “include” or “unclear” will be considered for the second phase (i.e. full text review). The decisions and reasons for exclusion will be recorded in Covidence software.

### Data extraction

Two research team members will independently perform data extraction using a form in Excel that is based on the JBI data extraction form for review of systematic reviews[39]. The data extraction form is provided as S2 file. We will conduct a pilot data extraction on 10% of eligible SRs until a consensus on extracted data is reached between reviewers. Any discrepancies will be resolved through discussion, or by consultation with a third reviewer. We will not extract data from the primary studies included in the reviews, nor re-synthetize their findings. Extracted information will include:

1. Review characteristics: first author, year of publication, country, type of review with or without meta-analysis; aim of the study; publication language; published protocol; number of databases searched, range date of search strategy; any restrictions (e.g. language, geographic or date), inclusion/exclusion criteria;
2. Participants: number and profile of participants (e.g patients, OHCPs);
3. Intervention and comparators: intervention(s) of interest and comparators; domains in dentistry; reported definition of intervention from authors;
4. Outcomes: primary outcomes (access to oral health care, patient -reported outcomes, patient reported-experiences); secondary outcomes relevant to patients and OHCPs; effect size metric(s) reported (e.g. risk ratio) for categorical outcomes and (e.g., standard mean differences) for continuous outcomes;
5. Setting: type of settings likely community, dental office, university, hospital;
6. Methods: type of study designs included (e.g. randomized controlled trials, observational studies or both); number of included studies, number of studies reporting data for meta-analyses, use of theoretical framework in the intervention or for data analysis; main primary outcomes of interest; risk of bias tools; statistical methods used to combine studies; synthesis and summary of data, estimates including heterogeneity measures, and any additional analyses (e.g. subgroup analysis or sensitivity analysis); the level of evidence, for instance the Grading of Recommendations, Assessment, Development and Evaluation (GRADE) from included studies;
7. Other details: main conclusions, limitations, next steps, funding source and conflicts of interest.

If we have missing data, we will search for additional information such as protocols or contact study authors to find the available information.

### Quality assessment of reviews

The quality assessment of SRs is an essential component when we conduct an overview of systematic reviews [32]. It includes both the methodological quality and the risk of bias which are distinct concepts [45, 46]. The quality assessment examines compliance with the highest possible standards in conducting and reporting their research process [47, 48], while risk of bias assessment (the appraisal of internal validity) examines any concerns with the design, conduct, analysis, interpretation, or reporting of a study which could affect the study’s results [49].

To conduct the quality assessment, we will use two valid and reliable tools covering complementary criteria: i) the AMSTAR-2 checklist (the Assessment of Multiple Systematic Reviews 2) to evaluate the methodological quality [50]; and ii) the ROBIS tool to assess comprehensively the risk of bias [51]. AMSTAR-2 is applicable to systematic reviews of both randomized controlled trials (RCTs) and non-randomized controlled trials (non-RCTs). This tool consists of 16 items, with 7 critical domains (items 2, 4, 7, 9, 11, 13, and 15). The assessment comprises three options, “yes” (item/question fully addressed), “no” (item/question not addressed), or “partial” (item/question not fully addressed). The overall confidence on the results of the review is classified into high, moderate, low, and critically low [50].

The ROBIS tool aims to assess the risk of bias in reviews related to interventions, etiology, diagnosis and prognosis [51]. It consists of 24 questions across three phases: Phase 1 assesses the relevance (optional); phase 2 identifies concerns with the review process; and phase 3 judges the risk of bias in the review. Phase 2 and 3 questions are answered with the following options: yes; probably yes; probably no; no; or no information. The concerns regarding phase 2 and phase 3 domains are classified as high, low, or unclear. We will focus only on phases 2 and 3 to give an overall score on the risk of bias in each review. Phase 2 identifies the concerns in the review across four domains: study eligibility criteria; identification and selection of studies; data collection and study appraisal; and synthesis and findings. There are signaling questions and a judgment of concerns about risk of bias for each domain (low, high or unclear). Phase 3 summarizes the concerns identified during the Phase 2 assessment [51]. Finally, we will provide a judgment regarding the overall risk of bias.

Despite the relevance of these tools and the growing number of reporting guidelines, they are mostly for SRs of quantitative reviews and there is a lack of critical appraisal tools [52] to assess the quality of SRs of qualitative studies [53]. In fact, some criteria from AMSTAR-2 and ROBIS tools are not adapted for quality assessment of factors such as risk of bias, publication bias, heterogeneity, meta-analysis in SRs of qualitative research [53]. However, we will use both these tools to conduct the quality assessment within the limit of their utilization.

Two reviewers will independently assess each review using both tools. Any discrepancies will be resolved by discussion or a consultation with a third reviewer. We will not perform any quality assessment of primary studies in included reviews.

### Data synthesis

We will perform a narrative synthesis of the data. We will present results in tabular form in tables describing characteristics of included studies (e.g. first author’s name, language of publication, country, settings, year of publication, profile of participants, study purpose), information on teledentistry (e.g. definition, teledentistry modalities), methods (e.g. SR with or without meta-analysis, type of analysis), additional results (e.g. assessment of quality, appraisal tool used, heterogeneity of the results, and level of evidence), and outcomes (e.g. primary outcomes and secondary outcomes from included studies). We will categorize the SRs into sub-groups, according to the type of intervention such as teledentistry modalities. If possible, we will perform a narrative synthesis of the subgroups.

An important concern in conducting an overview is the likelihood of overlap in primary studies across included reviews, which may result in overestimates in results (36) and confuse clinicians making decisions amongst competing interventions in their clinical practices. Thus, we will summarize key details from the studies included and will not perform a meta-synthesis of included meta-analyses. We will report any overlap between SRs in the tables as a matrix.

We will report the overall score of the two measurement tools (AMSTAR-2 and ROBIS), the level of evidence from the credibility assessment, and the percentage overlap between primary studies within included SRs and SR-meta-analysis (SR-MAs).

We will assess the certainty of evidence defined as any of evaluation of the totality or strength of the evidence on the impact of teledentistry using the following criteria for credibility assessment as proposed in four categories [38, 54]: Class I (Convincing evidence): associations with a statistical significance of P-value < 10^−6^, include more than 1000 cases (or more than 20 000 participants for continuous outcomes), have the largest component study reporting a significant result (P < 0.05), have a 95% prediction interval that excludes the null, does not have large heterogeneity (I² < 50%), and shows no evidence of small study effects (P > 0.10) and excess significance bias (P > 0.10); Class II (Highly suggestive evidence): associations with a significance of P < 0.001, include more than 1000 cases (or more than 20 000 participants for continuous outcomes), and have the largest component study reporting a statistically significant result (P < 0.05); Class III (Suggestive evidence): associations that report a significance of P < 0.01 with more than 1000 cases (or more than 20000 participants for continuous outcomes); Class IV (Weak evidence): remaining significant associations with P < 0.05. If we do not have sufficient data, we will analyse the certainty of evidence using data reported in the included SRs.

## Discussion

This review will contribute to a comprehensive body of knowledge on the potential effectiveness of teledentistry. To our knowledge, this overview is the first concerning teledentistry that will include a broad definition of teledentistry incorporating various modalities, participants, study designs, types of SRs and a combination of evidence, thereby increasing the understanding of the potential of teledentistry. This overview has implications for dental practice, policy, education and research. Beyond using a broad definition of teledentistry, this review will provide valuable knowledge on the use of digital technologies, including teledentistry, to improve oral health care systems. Teledentistry can be used to enhance the quality of oral health care as well as enhance oral health equity. The lack of high-quality information on the effectiveness of teledentistry has often been reported as a strong barrier to its implementation. This review has the potential to close that gap and will contribute to inform decision-makers, researchers, clinicians, and patients on the effectiveness of teledentistry for the delivery of care and to improve patients’ outcomes and experiences, as well as inform the needs for the future research. For instance, results will highlight teledentistry’s benefits and challenges, thereby identifying where is could potentially be used. These findings may also support the development and implementation of guidelines on teledentistry in clinical practice.

We anticipate some limitations. A first limitation relates to the overlap between the primary studies included in more than one SR in our overview, which can cause an overestimation of the effects for a given outcome. We will report the percentage of overlap between the included studies and will discuss of the impact of the overlap on our results. The second limitation is the restriction of our overview to SRs only, which could result in missing some relevant primary studies published during the completion of our review. To mitigate this issue, we will repeat our search just before we finish our manuscript and will identify any additional relevant published SRs, but we will only include them if they fulfill our inclusion criteria. We will document and report any deviations to the protocol during the review. In addition, we will mention any limitation in the lack of available critical appraisal tool for any designs (quantitative, qualitative and mixed reviews) [53].

## Dissemination plan

Results of this overview will be disseminated through presentations at national and international scientific and professional conferences, publication in peer-review journals and social media, such as Twitter, plus the website of the Faculty of Dental Medicine and Oral health Sciences, at McGill University.

## Data Availability

N/A

## Acknowledgement

The authors thank the summer students for their involvement in the first steps of data screening.

## Supporting information

S1 checklist: PRISMA-P (Preferred Reporting Items for Systematic review and Meta-Analysis Protocols) 2015 checklist: Recommended items to address in a systematic review protocol.

S1 file: Search strategy

S2 file: Data extraction sheet

## Funding

None

## Competing interests

None

## Availability of data and materials

Not applicable

## Notes

### Competing Interest Statement

The authors have declared no competing interest.

### Funding Statement

The author(s) received no specific funding for this work.

